# The willingness to accept and to pay for non-communicable disease screenings in rural Bangladesh: A contingent valuation study

**DOI:** 10.1101/2024.09.29.24314572

**Authors:** Lucie Sabin, Sarker Ashraf Uddin Ahmed, Abdul Kuddus, Carina King, Joanna Morrison, Sanjit Kumar Shaha, Naveed Ahmed, Tasmin Nahar, Kishwar Azad, Edward Fottrell, Hassan Haghparast-Bidgoli, Ali Kiadaliri

## Abstract

**Background:** Bangladesh is experiencing a rapid increase in non-communicable diseases (NCDs). Early screening is essential to enable timely treatment. In Bangladesh, access to screening services for NCDs is often limited. This study aimed to elicit the willingness to accept (WTA) and the willingness to pay (WTP) for four screening packages for NCDs and their association with the respondents’ sociodemographic characteristics among rural communities in Bangladesh.

**Methods:** A cross-sectional survey using contingent valuation was conducted in six villages of Faridpur district, Bangladesh. The WTP was elicited using a double-bounded dichotomous choice and bidding game approach, with initial bid amounts based on local market prices. Logistic regression was used to identify factors associated with WTA and WTP, and a two-part model was employed to estimate mean WTP and its determinants. We used a double-limit dichotomous choice model with the respondents who took part in the bidding exercise.

**Results:** We found that 83% of the 346 respondents were willing to accept at least one screening package and among these 44% would be willing to pay to accept all screenings. The willingness to accept and pay varied largely by the type of screening being offered, with much lower proportions for obesity than for hypertension and diabetes. The results suggested that individuals from households with a member with an NCD were more willing to accept and pay for screening compared to those from households without an NCD. Married individuals in the highest wealth and expenditure tertiles were also more willing to pay than not-married individuals from middle- and low-wealth and expenditure tertiles. Sex and occupation influenced the amount individuals were willing to pay.

**Conclusions:** Our results encourage the development of screening interventions for NCDs in rural populations. Further research is needed to assess their feasibility, effectiveness and equity in Bangladesh on a larger scale.

## Introduction

Non-communicable diseases (NCDs) represent a major and growing burden for global public health and pose considerable challenges for healthcare systems and individuals [1,2]. Globally, NCDs are the main cause of disability and mortality, and their prevalence and incidence are increasing. In 2023, NCDs accounted for approximately 74% of global mortality, resulting in approximately 41 million deaths, with 80% of NCD-related mortality driven by four major NCDs: cardiovascular disease, cancer, diabetes, and respiratory diseases [3]. The NCD mortality rate is significantly higher in low- and middle-income countries (LMICs), with 77% of the global mortality occurring in LMICs in 2023 [3], making NCDs a barrier to reducing health disparities between countries [4].

Similar to many LMICs, Bangladesh is experiencing a rapid increase in NCDs due to a range of factors including ageing of the population, tobacco consumption, lack of physical activity, unhealthy diets, and swift urbanisation [5]. The prevalence of major NCDs is notably high, with a 26.2% prevalence of hypertension [6] and 12.5% of the overall population reporting diabetes [7]. NCDs contribute significantly to mortality rates in Bangladesh with 67% of total deaths in the country attributable to NCDs in 2017 [8]. This increase in the prevalence and incidence of NCDs puts a strain on the healthcare system and the population [9]. Individuals affected by NCDs face challenges of increased healthcare costs, reduced quality of life, and higher mortality risks [2].

Early diagnosis and screening are essential to enable timely treatment, prevent disease progression and avoid the risk of complications [10]. In Bangladesh, access to screening services for NCDs remains limited [11], particularly in rural areas [12,13]. As part of the National NCD action plan, NCD corners have been established in Upazila health complexes levels since 2012 to provide NCD screening and related services [14]. However, comprehensive screening programs for NCDs are lacking, and existing NCD corners suffer from shortages of trained healthcare workers and essential supplies and medication [13]. When services to diagnose and manage NCDs are available, access to them can be hindered by costs, time, crowded conditions and distance [15].

Understanding the population’s willingness to accept (WTA) and willingness to pay (WTP) for NCD screening interventions is essential to inform their design and ensure their effective implementation. Contingent valuation studies, such as WTP evaluations, provide valuable information about individuals’ preferences and priorities when it comes to healthcare services. Although a few studies have investigated WTA and WTP for NCD screening in LMICs [16– 20], evidence remains limited, warranting further investigation. This study therefore aimed to elicit the WTA and the WTP for four screening packages for NCDs, and their association with the respondents’ sociodemographic characteristics among rural communities in Bangladesh. By assessing the preferences of individuals towards NCD screening, this research provides evidence to guide the development of feasible and accessible screening programmes. By looking at socioeconomic status, we are also looking at how equitable different options are.

## Methods

### Study design and setting

A cross-sectional survey including a contingent valuation (CV) was conducted to elicit WTA, WTP and the socioeconomic, demographic, and health-related factors associated with four screening packages for NCDs in rural Bangladesh. A CV survey consists of directly asking respondents about their willingness to pay or accept compensation for particular goods or services to estimate the monetary value individuals place on these items (Klose, 1999). The four screening packages included blood glucose testing; blood pressure measurement; height-weight measurement; and all three screenings combined. Each package also included a free health education leaflet covering diabetes, hypertension, and obesity, as well as their prevention and management.

The survey was conducted in six villages within the intervention area of the DClare NCD mobilisers’ participatory and learning action group meetings, located in Saltha Upazila in the Faridpur district of Bangladesh. Saltha was selected based on the availability of field staff from the local community, ease of communication from the district Sadar and the field office in Boalmari, and the logistical support provided by the field office. Faridpur is a district of 2,052.86 km^2^ with a population of around 2.16 million and a mainly agrarian economy [21]. The literacy rate in Faridpur is slightly below the national average at 73.27% amongst men, and 71.08% amongst women. Most of the population is Bengali and about 92% are Muslim [21]. Results from our survey conducted in Faridpur in 2016 revealed that the prevalence of intermediate hyperglycaemia and diabetes was 17.2% and 8.9% among men, and 23.4% and 11.5% among women, respectively. Nearly half of the men and women aged 30 years and above in our study sample were classified as prehypertensive or hypertensive. Furthermore, in a population-based survey/cross-sectional survey, only 25% of individuals with high blood glucose were aware of this. 75% of people with diabetes reported having suboptimal control [22].

### Study population, sample size and sampling procedure

Household heads aged 30 or over and living in the study villages for at least six months were eligible to be interviewed. The list of households from a census conducted in 2018 for the DMagic trial in the control villages was used as the sampling frame, with an estimated eligible population of 4,000 [23]. Using Krejcie and Morgan’s sampling method [24], we calculated a sample size of 400 (this includes an additional 10% sample to cover a potentially low response rate) which we considered adequate for the selected CV approach [25,26].

We aimed to sample 65 households from each of the 6 villages. To achieve this, we calculated a sampling interval for each village by dividing the total number of households in the village by 65. Households were then systematically sampled using this interval. The selection process began by randomly selecting the first household, determined by spinning a bottle or pen on the ground at the village centre and selecting the household it pointed to. Subsequently, data collectors proceeded clockwise from the initial household, recruiting every ‘n’ household based on the calculated interval (e.g., every 3rd household if the interval is 3). Ultimately, we successfully recruited 387 respondents from the 6 villages.

### Questionnaire design

We adapted the questionnaire from Yeo and Shafie (2018) and our previous CV survey conducted in rural Bangladesh [28]. We made necessary modifications to the tool after testing them in the study area, focusing primarily on simplifying the questions to ensure they were easily understandable for the largely illiterate study population. In addition to the WTA and WTP, we collected socio-demographic and socio-economic characteristics of individuals and households (see the survey questionnaire in S1 File).

### Data collection

Data were collected from 25 June to the 29 July 2023 on tablets using ODK Collect, an open-source Android App, by one male and one female local field worker, both experienced with ODK and part of the DMagic survey team. A male supervisor with experience in survey methods was recruited to supervise them. HHB trained the survey manager who was in charge of providing logistical support and coordinating the data collection. Subsequently, the supervisor and data collectors underwent four days of training on the survey methods in the field office, led by the survey manager. This was followed by a pilot in a village near Boalmari, Faridpur, where the Centre for Health Research and Innovation (CHRI) field office is located.

Data collectors faced several challenges, such as the paddy planting and jute harvesting season, during which people were often not available at home. Sometimes participants were interviewed while removing fibres from jute plants on the bank of the rivers or canals. Data collectors had to revisit households because the heads of households were busy with agricultural work. Despite this, the response rate was 89% after the second and third visits. Some people were reluctant to talk due to mistrust towards local NGOs and private clinics, stemming from their activities in the community. Additionally, some people were uninterested in blood glucose testing, fearing they might be diagnosed with diabetes. Some also expected incentives and free screening facilities in exchange for participating in the interview.

### Dependent variables

We included two dependent variables: WTA screening and WTP for screening. To determine the WTA, we asked respondents if they would like to be screened for diabetes, hypertension, obesity or a combination of all three if it was available in their community. We analysed WTA according to positive responses to any of the four screening packages compared with negative responses to all packages. Similarly to Yeo and Shafie (2018), we used a double-bounded dichotomous choice and bidding game approach to elicit respondents’ WTP amounts for the four screening packages (i.e. each screening individually and all of them together). In this approach, respondents were presented with two or three bid amounts, followed by an open-ended question to determine their maximum WTP for health screening (Fig 1). Subsequent bid amounts were adjusted based on respondents’ previous answers; affirmative responses prompted an increase in the bid, while negative responses led to a decrease. Respondents consistently answering ‘yes’ were further probed with an open-ended question to ascertain their maximum WTP. Respondents unwilling to pay were asked to provide reasons for their refusal. Fig 1 illustrates the methodology adopted in our study.

**Figure 1.**
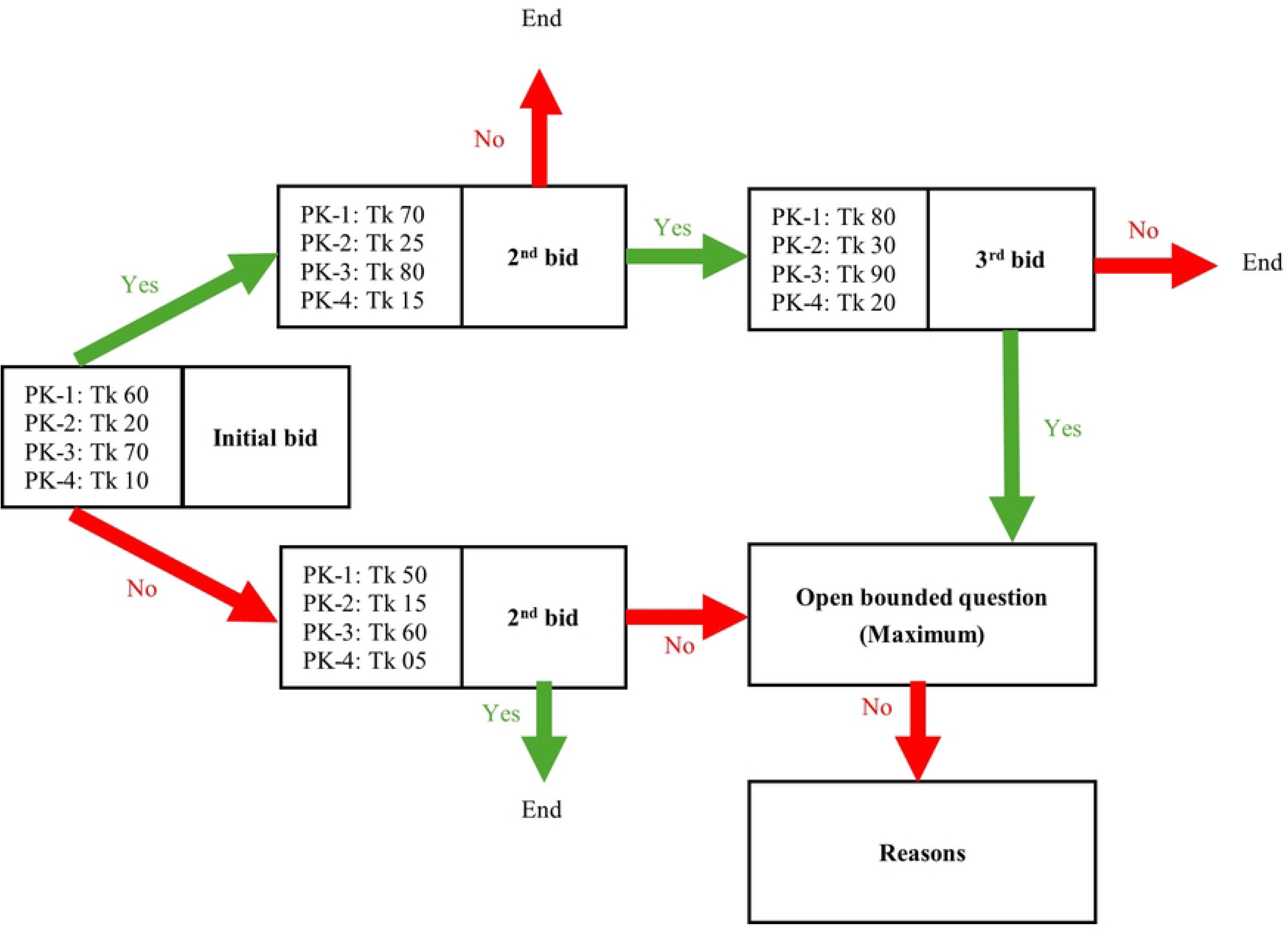
Double-bounded dichotomous choice and the bidding game approach. Notes: PK-1= blood glucose testing; PK-2=blood pressure measurement; PK-3=height-weight measurement; PK-4= all three screenings combined; Tk = Bangladeshi Taka

The initial bid amounts were determined based on the minimum market price of screening in Bangladesh. As government screening services are free of charge, the minimum screening prices for this intervention were estimated based on the prices of screening in local drug shops, pharmacies and private clinics. Travel costs to patients’ homes incurred by NCD mobilisers for screening visits were also considered. Similar to Yeo and Shafie (2018), we developed two sets of questionnaires (Fig 1) to mitigate potential bias related to the initial bidding, each featuring different initial bidding amounts for the screening packages. Respondents were randomly allocated to one of these sets.

WTP was measured in 2023 Bangladeshi Takas (Tk) and converted to US$ using the 2023 exchange rate (Tk 91.75=1US$) [29].

### Independent variables

We included sex, age, literacy, occupation (housework/housewife, farmer, other jobs, unemployed/not working), marital status, household wealth and household expenditure tertiles and the reported presence of NCDs (diabetes, high blood pressure, obesity or heart disease) in the household. Household wealth index was constructed using principal component analysis (PCA) incorporating data on ownership of assets, agricultural land, access to utilities and infrastructure, and housing characteristics. Household expenditure was measured using monthly spending on food and non-food items. Although household wealth and expenditure might be correlated, both were used as they measure different aspects of households’ social and economic status.

### Statistical analysis

The data was analysed using logistic regression to assess the associations between covariates and WTA (yes vs. no) and WTP (yes vs. no) for the screening alternatives. We estimated both unadjusted and adjusted logistic regressions.

We used a parametric two-part model to elicit the mean WTP value and the factors associated with the mean WTP [30,31]. To do so, we first determined the maximum WTP as the mid-point between the lower acceptance bid and the higher rejection bid [27]. For example, for a respondent answering “yes” to Tk 60 and “no” to Tk 70, we established Tk 65 as the maximum WTP. For respondents who consistently answered “yes” or “no” to the bid options, we considered their response to the open-ended question as the maximum WTP. We allocated a maximum WTP of zero for respondents who wished to accept the screening but indicated they did not wish to pay for it. In the two-part model modelling, we initially used a binary choice model to evaluate the probability of observing either a positive or zero outcome (WTP=0 or >0). In the second step, a linear regression model was applied, based on the positive results identified in the initial stage. This approach aimed to mitigate estimation biases by accommodating the heterogeneous densities of zero and non-zero values. We then used the *margins* command in Stata to estimate the marginal effect of each covariate on the WTP.

In addition to the two-part model modelling, we used a double-limit dichotomous choice model with the respondents who took part in the bidding exercise (i.e. excluding those who wished to accept the screening but indicated they did not wish to pay for it). Two bids and corresponding “yes/no” answers were modelled using *doubleb* command in Stata [32]. For respondents with 3 bids (i.e. those responding “yes” and “yes” to the first and second bids), we treated the second bid as the initial one.

All statistical analyses were performed in Stata 18, College Station, TX: StataCorp LLC.

### Ethics

We obtained ethical approval for this study, as part of the DClare project, from the University College London Research Ethics Committee (4199/007) and the Ethical Review Committee of the Diabetic Association of Bangladesh (BADAS-ERC/E/19/00276). All respondents gave informed written consent before taking part in the survey.

## Results

### Socio-demographic characteristics of respondents

Descriptive statistics for the sample are presented in Table 1. The total sample size was 387 with 41 non-respondents, resulting in 346 individuals considered for this analysis. As questionnaires were administered to household heads, most of the respondents were males (80%). The age distribution showed most respondents in the 30-49 age group (58%), male (81%) and married (92%). 34% of the sample was literate, 19% were doing housework and 47% were farmers.

**Table 1.** Descriptive statistics of the sampled respondents.

| **Variables** | **Frequencies** |
| --- | --- |
| Age |  |
| 30-49 years | 200 (57.8%) |
| 50-64 years | 99 (28.6%) |
| 65+ years | 45 (13.0%) |
| Missing | 2 (0.6) |
| Sex |  |
| Female | 67 (19.4%) |
| Male | 279 (80.6%) |
| Expenditure tertile |  |
| Lowest tertile | 116 (33.5%) |
| Middle tertile | 115 (33.2%) |
| Highest tertile | 115 (33.2%) |
| Household wealth tertile |  |
| Lowest tertile | 116 (33.5%) |
| Middle tertile | 115 (33.2%) |
| Highest tertile | 115 (33.2%) |
| Marital status |  |
| Married | 317 (91.6%) |
| Not married | 29 (8.4%) |
| Literate |  |
| Yes | 117 (33.8%) |
| No | 229 (66.2%) |
| Occupation |  |
| Housework/housewife | 64 (18.5%) |
| Farmer | 161 (46.5%) |
| Other jobs | 100 (28.9%) |
| Unemployed/not working | 21 (6.1%) |
| Presence of NCD in household |  |
| Yes | 248 (71.7%) |
| No | 98 (28.3%) |
| Total | 346 |

### Willingness to accept screening and associated factors

Overall, 287 (83.0%) out of 346 respondents said they wished to accept at least one screening package. Among these, 124 (43.2%) of respondents wished to accept all screening programs. Among those who reported that they would accept screening (n=287), when we looked at each specific package, 67% of respondents said that they wished to accept screening for diabetes, 62% for hypertension, and 39% for obesity. S4 Table presents the frequency of respondents’ willingness to accept screening for diabetes, hypertension or obesity by socio-economic characteristics. A higher proportion of men than women were willing to accept screening for all three conditions. Younger respondents, particularly those aged 30 to 49, showed the highest proportion of willingness to accept screening for all conditions, with this proportion decreasing slightly in older age groups. A higher proportion of people in the lowest wealth tertile were willing to be screened for obesity, while a higher proportion of people in the highest wealth tertile were willing to be screened for diabetes and hypertension. A higher proportion of farmers and married respondents were willing to accept screening.

Sociodemographic factors associated with WTA of any screening package are presented in Table 2. Among independent variables, having a household member with an NCD was the only variable that had a statistically significant association with WTA. Regarding the associations with other variables, the 95% CI were wide and inconclusive.

**Table 2.** Sociodemographic factors associated with the willingness to accept any screening.

|  | **Univariable** |  | **Multivariable** |  |
| --- | --- | --- | --- | --- |
|  | Odds Ratio | 95% CI | Odds Ratio | 95% CI |
| Male | 1.22 | 0.62, 2.41 | 1.16 | 0.43, 3.15 |
| Age |  |  |  |  |
| 30-49 | Ref. |  |  |  |
| 50-64 | 0.92 | 0.49, 1.73 | 0.66 | 0.33, 1.35 |
| 65+ | 1.11 | 0.46, 2.70 | 0.75 | 0.26, 2.12 |
| Household wealth tertile |  |  |  |  |
| Lowest | Ref. |  |  |  |
| Medium | 1.12 | 0.56, 2.21 | 0.95 | 0.45, 2.01 |
| Highest | 1.12 | 0.56, 2.21 | 0.73 | 0.32, 1.63 |
| Married | 0.76 | 0.26, 2.28 | - | - |
| Literate | 0.91 | 0.51, 1.64 | 0.84 | 0.43, 1.64 |
| Work status |  |  |  |  |
| Housework/housewife | Ref. |  |  |  |
| Farmer | 1.39 | 0.67, 2.86 | - | - |
| Other jobs | 1.59 | 0.71, 3.56 | - | - |
| Unemployed/not working | 1.68 | 0.43, 6.53 | - | - |
| Earner | 0.93 | 0.50, 1.74 | 1.01 | 0.40, 2.56 |
| Household expenditure tertile |  |  |  |  |
| Lowest | Ref. |  |  |  |
| Middle | 1.22 | 0.63, 2.33 | 1.08 | 0.52, 2.24 |
| Highest | 1.75 | 0.85, 3.59 | 1.60 | 0.72, 3.54 |
| Presence of NCD in household | 5.91 | 2.46, 14.19 | 6.62 | 2.67, 16.41 |

Notes: CI: Confidence Interval. Almost all males (98.2%) were married, hence the variable indicating the marital status was excluded from the analysis. Work status variable excluded from the multivariable analysis because it was strongly correlated with the sex variable.

### Willingness to pay for the screening and associated factors

Among those who wished to accept any screening package (n=287), 233 (83.0%) individuals wished to pay for at least one screening package, with 103 (44.2%) of these 233 individuals saying that they would be willing to pay to accept all screening packages. Among those willing to pay, when we looked at specific packages, 59% of respondents said that they would be willing to pay for diabetes screening, 55% for hypertension screening and 30% for obesity screening. S5 Table presents the frequency of respondents’ willingness to pay for screening for diabetes, hypertension or obesity by socio-economic characteristics. The proportions for willingness to pay according to socio-economic characteristics were similar to those observed for willingness to accept screening. Sociodemographic factors associated with the willingness to pay for screening are presented in Table 3. These individuals were, on average, more likely to be married, in the highest wealth tertile, in the highest expenditure tertile, and to have an NCD in the household.

**Table 3.**
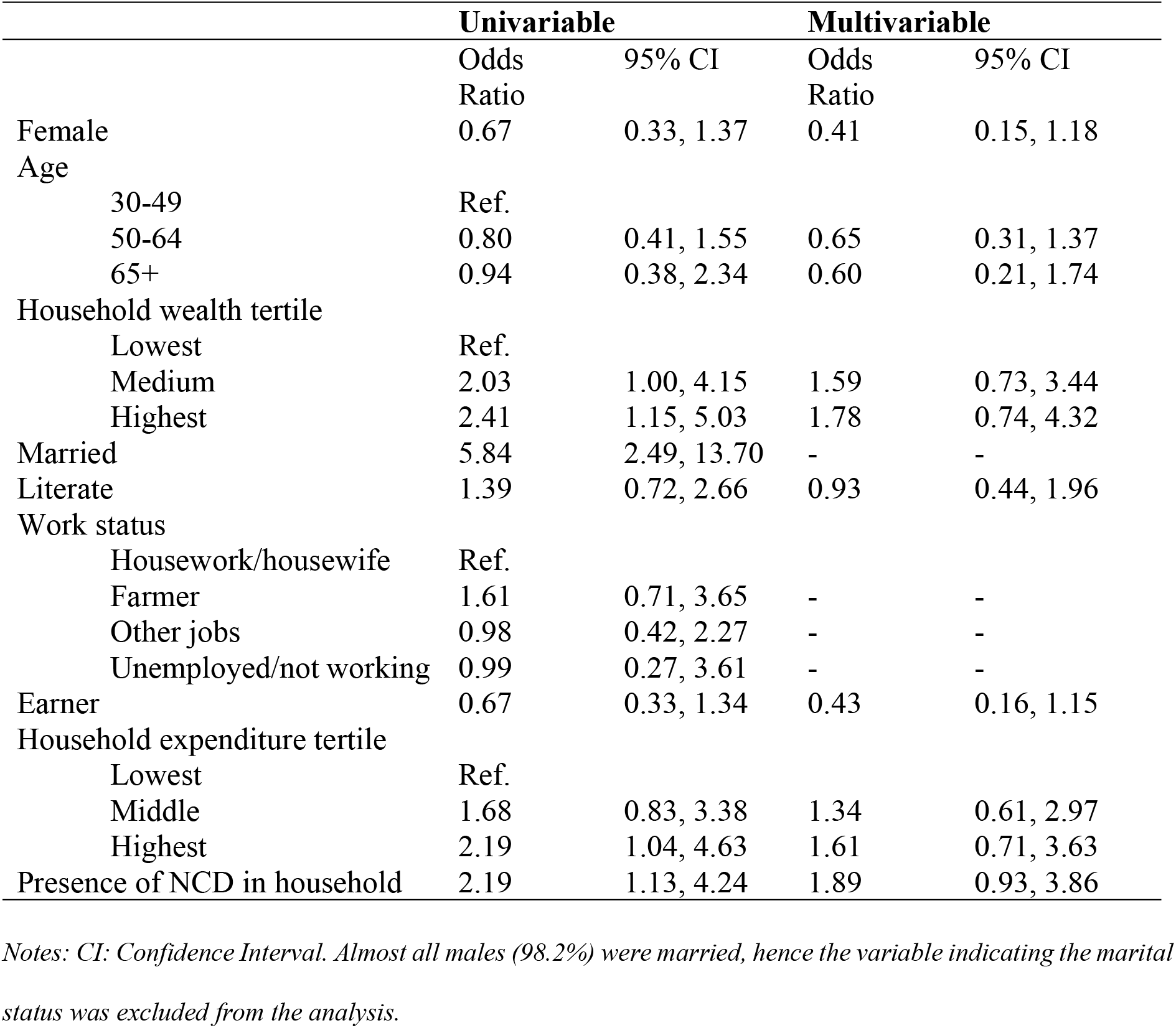
Associated factors with WTP for all screening defined as intention to pay an amount of money.

|  | Univariable |  | Multivariable |  |
| --- | --- | --- | --- | --- |
|  | Odds Ratio | 95% CI | Odds Ratio | 95% CI |
| Female | 0.67 | 0.33, 1.37 | 0.41 | 0.15, 1.18 |
| Age |  |  |  |  |
| 30-49 | Ref. |  |  |  |
| 50-64 | 0.80 | 0.41, 1.55 | 0.65 | 0.31, 1.37 |
| 65+ | 0.94 | 0.38, 2.34 | 0.60 | 0.21, 1.74 |
| Household wealth tertile |  |  |  |  |
| Lowest | Ref. |  |  |  |
| Medium | 2.03 | 1.00, 4.15 | 1.59 | 0.73, 3.44 |
| Highest | 2.41 | 1.15, 5.03 | 1.78 | 0.74, 4.32 |
| Married | 5.84 | 2.49, 13.70 | - | - |
| Literate | 1.39 | 0.72, 2.66 | 0.93 | 0.44, 1.96 |
| Work status |  |  |  |  |
| Housework/housewife | Ref. |  |  |  |
| Farmer | 1.61 | 0.71, 3.65 | - | - |
| Other jobs | 0.98 | 0.42, 2.27 | - | - |
| Unemployed/not working | 0.99 | 0.27, 3.61 | - | - |
| Earner | 0.67 | 0.33, 1.34 | 0.43 | 0.16, 1.15 |
| Household expenditure tertile |  |  |  |  |
| Lowest | Ref. |  |  |  |
| Middle | 1.68 | 0.83, 3.38 | 1.34 | 0.61, 2.97 |
| Highest | 2.19 | 1.04, 4.63 | 1.61 | 0.71, 3.63 |
| Presence of NCD in household | 2.19 | 1.13, 4.24 | 1.89 | 0.93, 3.86 |

Notes: CI: Confidence Interval. Almost all males (98.2%) were married, hence the variable indicating the marital status was excluded from the analysis.

### The amount of willingness to pay

The average unadjusted WTP value for all screenings was 41.2 Bangladeshi Taka (0.45 US$; 95% CI 36.4, 46.0). It was Tk 9 (0.098 US$; 95% CI 7.7, 10.3) for screening for hypertension, Tk 27.2 (0.30 US$; 95% CI 24.1, 30.3) for diabetes and Tk 6.6 (0.072 US$; 95% CI 5.3, 7.9) for obesity. Table 4 presents the results from the double-bounded and two-part models. When considering all screening, there were no conclusive differences in the amount of WTP according to the respondents’ sociodemographic characteristics.

**Table 4.**
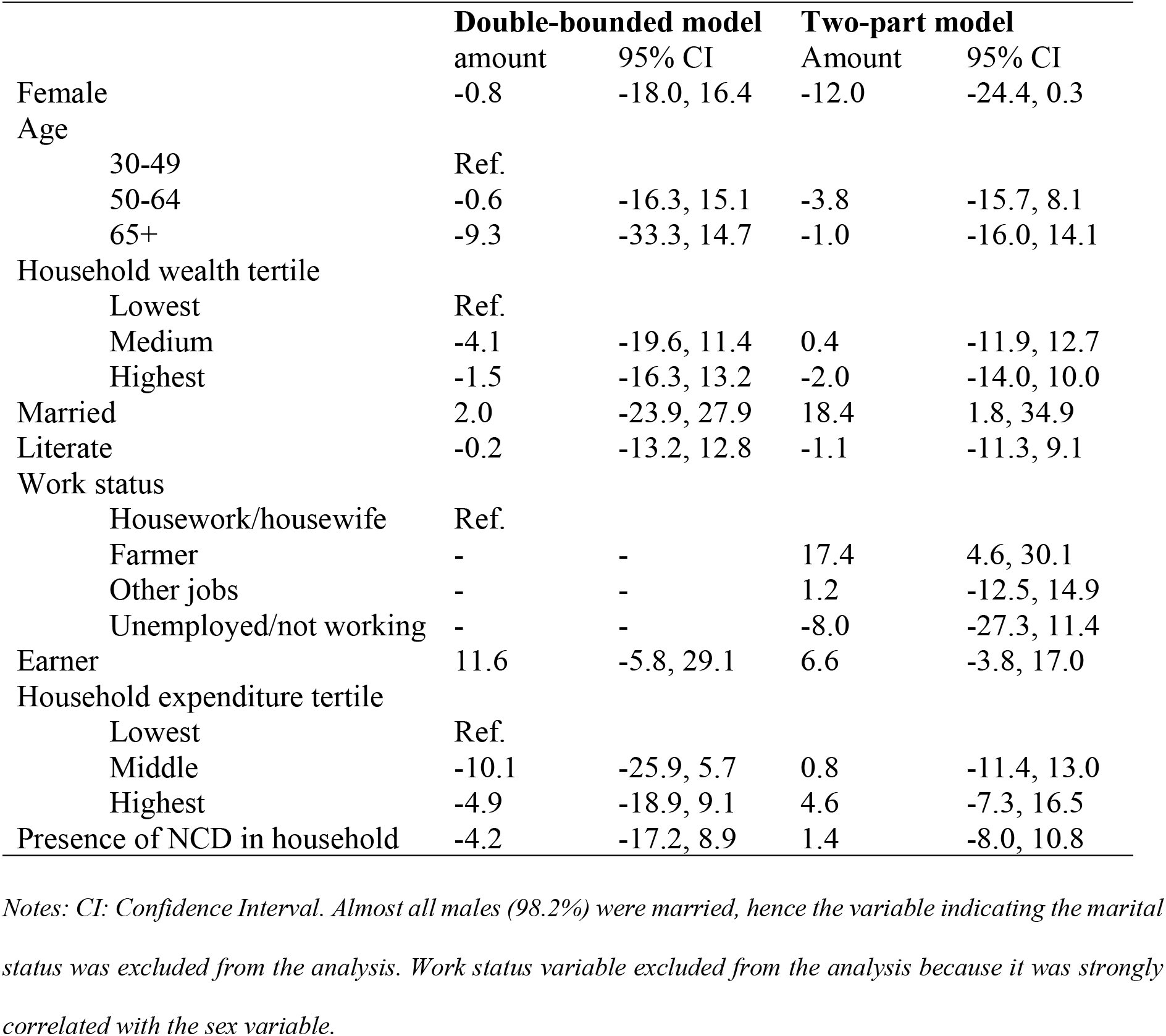
Willingness-to-pay estimates from double-bounded and two-part unadjusted models for all screening.

The two-part model indicated that women were willing to pay Tk 12 (95% CI -24.4, 0.3) less than men. Married individuals were willing to pay, on average, around Tk 18.4 (1.8, 34.9) more than unmarried people. On the other hand, people who had a job other than farming or housework were willing to pay a higher amount than other people.

Results from the double-bounded and two-part models for diabetes, hypertension, and obesity are presented in S1, S2 and S3 Tables. The results are generally similar to those of the main analysis, although the 95% CIs are narrower for some variables.

## Discussion

In the current study, we elicited the WTA and the WTP for screening for NCDs in rural Bangladesh. The results suggested that individuals in households where a household member has an NCD were more willing to accept and pay for screening than in households where no members had an NCD. Married individuals in the highest wealth and expenditure tertiles were also more willing to pay for screening than single individuals from middle- and low-wealth and expenditure tertiles. Gender and occupation influenced the amount people were willing to pay.

We found that 83% of respondents were willing to accept at least one of the screening packages. A qualitative study conducted in Bangladesh [17], examined WTA services for NCDs from primary health facilities and identified knowledge and perceived need for NCD care as key factors influencing WTA. Similarly, we found that individuals from households with experience of an NCD were more willing to accept screening than those without an NCD, which was possibly due to their higher risk perception and consideration of screening importance.

Among those who wished to accept any screening package, we found that 44% of respondents would be willing to pay to accept all screenings. In studies conducted in Taiwan [18] and China [19], similar results were found with respectively 65% and 60% of respondents willing to pay for diabetes screening compared with 59% of respondents in our study. However, the proportion of those with WTP for hypertension screening found in Bulgaria was much higher (95%) [20]. This might be explained by the sample being more health-conscious in the study conducted in Bulgaria as they were sampled from a community pharmacy and almost half of them declared measuring their blood pressure once a month. In addition, strong public health initiatives and a high level of awareness of cardiovascular diseases, given the high mortality rate from these diseases in Bulgaria, may contribute to this higher WTP [33].

We found that the proportion of respondents with the willingness to accept and pay varied largely by the type of screening being offered, with much lower proportions for obesity than for hypertension and diabetes. This difference could be because obesity is much easier to subjectively and objectively measure than diabetes and hypertension. Additionally, although in some cultures being overweight is desirable and can be seen as a proxy for wealth and sexual attractiveness, in Bangladesh, obesity is the subject of stigma and a study found an association between obesity and depression, anxiety, and stress [34]. The stigma of obesity could reduce the WTA and WTP for screening, as people may feel too embarrassed or ashamed to be screened.

The amount that respondents were willing to pay for all screenings reported in the present study (0.45 US$) varied largely by disease (0.098 US$ for hypertension, 0.30 US$ for diabetes and 0.072 US$ for obesity). It was much lower than the WTP amount found in previous studies conducted in Taiwan (14.3 US$) and China (1.6 US$ to 4.7 US$) for diabetes screening [18,19] and in Bulgaria for hypertension screening (1.30 US$) [20]. These differences can be attributed to disparities in purchasing power between countries, with GDP per capita being much lower for Bangladesh (2,688.3 US$ per capita) than for China (12,720.2 US$ per capita) or Bulgaria (13,974 US$ per capita) [35].

Socioeconomic status of individuals was one of the main predictors of the WTP for screening with people in the highest asset and expense tertiles being more willing to pay for screening. Opposite findings were found in China with people from lower-income households being more likely to be willing to pay. We also found that people from the middle and highest tertiles for expenditure and wealth were willing to pay more for screening for NCDs. Similarly to Xiao et al. (2023), we found that females were willing to pay lower sums than males. T This aligns with the context of Bangladesh, where women have less control over expenditure in the households [36]. We also found that people from households with a member with an NCD were willing to pay more. This is consistent with the results from a study in Taiwan with people with severe stages of diabetic retinopathy being willing to pay a higher amount for screening than people with mild or no retinopathy [18]. In Bulgaria, individuals who measured their blood pressure every day or once a week were willing to pay more than the ones who did it occasionally [20].

Our results suggest that poorer people are less willing to pay than better-off people suggesting the importance of addressing affordability barriers to ensure accessibility and affordability of screening services for all segments of the population. The results also suggest the importance of pairing screening packages with public health interventions to increase awareness of the importance of early detection for a positive and less costly outcome [15].

Our study has some limitations. First, although we trained data collectors to avoid bias, data collection relied on self-reported responses, which may be subject to social desirability bias that can affect the accuracy of reported acceptance and WTP. Second, as Bangladesh is a collective society where individuals do not make financial decisions alone, the use of open-ended contingent valuation methods to obtain WTP may introduce variability in responses. Thirdly, our sample consisted mainly of heads of household, most of whom were men, which limits the representation of women’s willingness to pay (WTP) and willingness to respond (WTA). This should be taken into account when interpreting the results, as women often have limited control over household resources and are dependent on men for financial decisions. In this regard, women’s views on household money management may differ from those of men. Lastly, the small sample size of our study resulted in wide 95% confidence intervals, highlighting the need for larger samples in future research to improve the accuracy and reliability of the results.

These limitations aside, our results advance the empirical literature on WTA and WTP for NCD screening in LMICs. To the best of our knowledge, this study is the first to have been carried out on a general population, community-based sample in rural areas. Our results suggest a relatively high willingness to accept screening, particularly among respondents who are more aware of non-communicable diseases.

## Conclusion

High incidence and mortality from NCDs in Bangladesh and LMICs in general indicate the need for low-cost prevention interventions including screening. This study showed a relatively strong acceptance to accept screening, particularly among those already with experience of NCDs in their household. Our results are encouraging for the development of screening interventions for NCDs, and further research is needed to assess their feasibility, effectiveness, equity and scalability in Bangladesh on a larger scale. Particular attention should be paid to designing interventions to ensure inclusion in screening campaigns.

## Data Availability

Anonymised data can be shared upon request from the authors.

## Author Contributions

Study design; HH-B, Aki, SAUA. Data collection; SAUA, SKS, AKu, NA, KAk, TN, KAz. Data analysis; AKi, with contribution from HH-B, and LS. Manuscript draft writing; LS, with support from HH-B and AKi. Critical review and edit; HH-B, EF, JM, CK, and AKi. Primary supervision; HH-B. All authors have commented, read and approved the final version of the manuscript.

